# Mortality in COVID-19 amongst women on Hormone Replacement Therapy or Combined Oral Contraception: A cohort study

**DOI:** 10.1101/2021.02.16.21251853

**Authors:** Hajira Dambha-Miller, William Hinton, Mark Joy, Michael Feher, Simon de Lusignan

## Abstract

**Objective:** To investigate the association between Hormone Replacement Therapy (HRT) or Combined Oral Contraception (COCP) use, and the likelihood of death in women with COVID-19.

**Design:** A cohort study

**Setting:** 465 general practices in England within the Oxford-Royal College of General Practitioners (RCGP) Research and Surveillance Centre (RSC) primary care database.

**Population:** 1,863,478 women aged over 18 years

**Methods:** We identified a cohort of women with COVID-19 from the computerised medical records of the RCGP RSC database. Mixed-effects logistic regression models were used to quantify the association between HRT or COCP use, and all-cause mortality among women with COVID-19 in unadjusted and adjusted models.

**Results:** There were 5451 COVID-19 cases within the cohort. HRT was associated with a significantly lower likelihood of all-cause mortality in COVID-19 (adjusted OR 0.22, 95%□CI 0.05 to 0.94). There were no reported events for all-cause mortality in women prescribed COCPs. This prevented further examination of the impact of COCP.

**Conclusions:** Women on HRT with COVID-19 had a lower likelihood of death. Further work is needed in larger cohorts to examine the association of COCP in COVID-19. Our findings support the current hypothesis that oestrogens may contribute a protective effect against COVID-19 severity.

**Funding:** This study was funded by a School for Primary Care National Institute for Health Research grant (SPCR2014-10043).

## INTRODUCTION

The novel severe acute respiratory syndrome coronavirus 2 (SARS-CoV-2) continues to spread globally with males and females equally susceptible to the infection. However, males experience greater severity of infection with higher rates of hospitalisation and mortality.[1] A recent review of sex differences in COVID-19 including data from 38 countries reported mortality in males as 1.7 times higher than the average female. [2] Similar data have been observed in previous pandemics including the SARS-CoV (Severe Acute Respiratory Syndrome Corona Virus) and MERS-CoV (Middle East Respiratory Syndrome Corona Virus) outbreaks.[3] The reason for these sex differences is unclear. A range of hypotheses have been proposed from variations in patterned sex behaviours such as smoking, co-morbidities and sex-based immunological variations.[2] In particular, the role of oestrogen in female immune responses has received much attention.[4,5] Younger females or those with higher oestrogen levels are less likely to experience severe COVID-19 complications.[4] Earlier studies show that females mount faster and greater immune responses to viral infections through cellular and humoral immune responses.[2] Moreover, immune responses can be modulated by oestrogen through a reduction in T-cell exhaustion and suppression of IL-1β and IL-6 production.[6] This potentially limits the cytokine storm and subsequent respiratory failure that is characteristically triggered by SARS-CoV-2. This may explain why fewer women compared to men have been hospitalised, admitted to ITU or have died during the pandemic. [1]

Recent observational data suggests that women aged 18-45 years taking the COCP have a lower risk of COVID-19 infection (P=<0.001) and a lower rate of hospital attendance (P=0.023). [7,8] Evidence on Hormone Replacement Therapy (HRT) has been more inconsistent.[7] To our knowledge, no studies have investigated the role of oestrogen containing products on mortality in COVID-19. The potential protective effects of oestrogen on the severity of COVID-19 has important public health and clinical relevance. In the absence of curative treatment for the infection, repurposing of existing drugs including exogenous oestrogen products requires rapid investigation. It is also necessary to understand the potential impact of these drugs for women taking them given public and prescriber concern. Accordingly, we quantified the association between COCP or HRT use, and the likelihood of mortality amongst females with COVID-19 during the first six months of the pandemic.

## METHODS

### Study design, data source and population

In this retrospective cohort study, we used the Oxford-Royal College of General Practitioners (RCGP) Research and Surveillance Centre (RSC) database of individual-level pseudonymised data that has been routinely collected from primary care records.[9] It includes continuous longitudinal data with sociodemographic information, prescribed medications, clinical diagnosis, symptoms, investigations and results. The database includes 465 GP practices in both rural and urban areas of England covering a nationally representative population of 3.7 million people. Within the database, we identified a cohort of women registered on the 1st January 2020 who were aged over 18 years with confirmed or probable COVID-19. Confirmed cases were defined as those with a positive RT-PCR assay for SARS-CoV-2 on a nasal or pharyngeal swab and probable cases were those diagnosed radiologically or clinically based Public Health England’s recommendations. Clinical symptoms included a new continuous cough, a fever >37.8 degrees or a loss/change in normal smell or taste. The variability in the availability of RT-PCR testing during the pandemic meant that most recorded cases in the dataset were diagnosed as probably cases. [10,11] Our previous work shows that clinical and probable cases are similar in terms of outcomes; for mortality, the odds ratios were 8.9 (95% CI = 6.7 to 11.8, *P*<0.0001) and 9.7 (95% CI = 7.1 to 13.2, *P*<0.0001) for RT-PCR confirmed and clinically diagnosed cases, respectively. [12]

### Exposure: HRT or COCP use

We defined the exposure as one or more HRT or COCP prescriptions within 6 months of a confirmed or suspected COVID-19 case. This had to be before case confirmation.

### Outcome: All-cause mortality

The primary outcome was all-cause mortality during the follow-up period (1st January to 21st June 2020) as recorded in the electronic record.

### Covariables

We extracted data on age, ethnicity and socioeconomic status. Ethnicity was self-reported in the records.[13] For socioeconomic status, the English Index of Multiple Deprivation was used. [14,15] We combined IMD quintiles 1 and 2 because recent evidence shows that there is a low frequency of testing, leading to sparse data in the most deprived quintile.[14] We included the most recently available data on the household size as this is important in acquiring COVID-19 infection.[16] For clinical variables, we considered body mass index (BMI) as the most recent recording within the 12 months before the study start date and coding for co-morbidities were recorded as any history of hypertension, coronary heart disease, type 1 diabetes, type 2 diabetes, asthma, chronic obstructive pulmonary disease (COPD), and chronic kidney disease (CKD) stage 3-5. Smoking status categorised as non-smoker, active-smoker or ex-smoker. We also included prescriptions for prednisolone and/or disease-modifying anti-rheumatic drugs as a surrogate for immunosuppression. We used standardised coding required for NHS payment and administrative purposes to increase consistency and quality of data included.

### Statistical Analysis

Sociodemographic and clinical characteristics were summarised using descriptive statistics, and we compared the characteristics of those with and without missing data. Univariable logistic regression models were used to quantify the association between HRT and COCP (separately) in relation to all-cause mortality. We then ran multivariable models adjusting for age, sex, ethnicity, index of multiple deprivation, household size, BMI, and comorbidities. Mixed-effects models were performed to account for practice clustering. We ran a complete case analysis. Statistical analyses were performed using R (Version 3.5.3). The level of significance was set at 5% and all statistical tests were 2-tailed. Model parameters were reported using odds ratios (ORs) and 95% confidence intervals. Our findings are reported in line with the STROBE and RECORD guidelines for observational studies using routinely collected health data.

### Patient and public involvement

Patients and members of the public contributed to the setting the research question, the outcome measures and the dissemination of our findings.

### Ethical approval

This study received approval from the Oxford-RCGP RSC study approval committee (RSC_0920) and the University of Southampton Research Ethics committee (56309)

## RESULTS

### Participant characteristics

In this retrospective cohort study, the denominator population included 1,863,478 women across 465 general practices within the Oxford-RCGP RSC database during the first six months of the UK’s COVID-19 pandemic. Within this sample, we identified a cohort of 5451 women who had COVID-19. The mean follow-up period was 164.9(SD 19.6) days. The mean age of the cohort was 59.0 years (SD 21.7); self-assigned ethnicity was predominantly White (64.8%). There were 235 women with HRT prescriptions and 171 with a prescription for the COCP. Table 1 summarises sociodemographic and clinical characteristics in the whole cohort, and separated as those on HRT or COCP. During the follow up-period 664 (12.2%) women died. Table 2 summarises the characteristics of women who died; they were more likely to be older with multiple morbidities.

**Table 1:** Baseline characteristics of women with COVID-19 in the RCGP RSC database presented by those on HRT, COCP or neither drug.

|  | **Total**<br>(N= 5451) | **Neither drug**<br>(N = 5045) | **HRT**<br>(N = 235) | **COCP**<br>(N = 171) |
| --- | --- | --- | --- | --- |
| ***Sociodemographic*** |  |  |  |  |
| Age (years) * | 59.0 (21.7) | 60.2 (21.7) | 54.6 (9.4) | 29.3 (7.4) |
| Ethnicity recorded | 4356 (79.9) | 4024 (79.8) | 193 (82.1) | 139 (81.3) |
| White | 3534 (64.8) | 3231 (64.0) | 179 (76.2) | 124(72.5) |
| Asian | 510 (9.4) | 497 (9.9) | 6 (2.6) | 7 (4.1) |
| Black | 211 (3.9) | 203 (4.0) | 4 (1.7) | 4 (2.3) |
| Mixed & other | 101 (1.9) | 93 (1.8) | 4 (1.7) | 4 (2.3) |
| IMD Quintile recorded | 5326 (97.7) | 4931 (97.7) | 232 (98.7) | 163 (95.3) |
| 5 (Least deprived) | 1136 (20.8) | 1018 (20.2) | 74 (31.5) | 44 (25.7) |
| 4 | 1088 (20.0) | 999 (19.8) | 58 (24.7) | 31 (18.1) |
| 3 | 1054 (19.3) | 986 (19.5) | 36 (15.3) | 32 (18.7) |
| 1&2 (Most deprived) | 2048 (37.6) | 1928 (38.2) | 64 (27.2) | 56 (32.7) |
| Settlement or population density | 5328 (97.7) | 4933 (97.8) | 232 (98.7) | 163 (95.3) |
| Rural | 933 (17.1) | 833 (16.5) | 65 (27.7) | 35 (20.5) |
| Urban | 4395 (80.6) | 4100 (81.3) | 167 (71.1) | 128 (74.9) |
| ***Clinical*** |  |  |  |  |
| BMI recorded | 5122 (94.0) | 4724 (93.6) | 231 (98.3) | 167 (97.7) |
| BMI (kg/m<sup>2</sup>) | 28.2 (7.3) | 28.3 (7.3) | 29.3 (6.4) | 24.55 (4.5) |
| Smoking status recorded | 5328 (97.7) | 4928 (97.7) | 233 (99.1) | 167 (97.7) |
| Non-smoker | 2128 (39.0) | 1969 (39.0) | 67 (28.5) | 92 (53.8) |
| Active-smoker | 486 (8.9) | 447 (8.9) | 28 (11.9) | 11 (6.4) |
| Ex-smoker | 2714 (49.8) | 2512 (49.8) | 138 (58.7) | 64 (37.4) |
| Co-morbidity* | 3001 (55.1) | 2859 (56.7) | 116 (49.4) | 26 (15.2) |
| All medications** | 2452 (45.0) | 2333 (46.2) | 100 (42.6) | 19 (11.1) |

\* Includes hypertension, coronary heart disease, diabetes (type 1 or type 2), chronic kidney disease stage 3-5, asthma, COPD, immunocompromised
\*\* Includes antihypertensive medication, lipid-lowering medication, hypoglycaemic medication, inhalers, immunosuppressants

**Table 2:** Baseline characteristics of women with COVID-19 who died during the follow-up period in the Oxford-RCGP RSC database.

|  | **Total**<br>N= 5451 | **Non-decedent**<br>N=4787 | **Decedent**<br>N=664 |
| --- | --- | --- | --- |
| Age (years) | 59.0 (21.7) | 55.7 (20.8) | 82.5 (11.3) |
| Ethnicity recorded | 4356 (79.9) | 3838 (80.2) | 518 (78.0) |
| White | 3534 (64.8) | 3064 (64.0) | 470 (70.8) |
| Asian | 510 (9.4) | 484 (10.1) | 26 (3.9) |
| Black | 211 (3.9) | 200 (4.2) | 11 (1.7) |
| Mixed other | 101 (1.9) | 90 (1.9) | 11 (1.7) |
| IMD Quintile recorded | 5326 (97.7) | 4671 (97.6) | 655 (98.6) |
| 5 (Least deprived) | 1136 (20.8) | 993 (20.7) | 143 (21.5) |
| 4 | 1088 (20.0) | 936 (19.6) | 152 (22.9) |
| 3 | 1054 (19.3) | 918 (19.2) | 136 (20.5) |
| 1&2 (Most deprived) | 2048 (37.6) | 1824 (38.1) | 224 (33.7) |
| Settlement or population density | 5328 (97.7) | 4671 (97.6) | 657 (98.9) |
| Rural | 933 (17.1) | 816 (17.0) | 117 (17.6) |
| Urban | 4395 (80.6) | 3855 (80.5) | 540 (81.3) |
| BMI recorded | 5122 (94.0) |  |  |
| BMI (kg/m<sup>2</sup>) | 28.2 (7.3) | 28.4 (7.3) | 26.6 (7.1) |
| Smoking status recorded | 5328 (97.7) | 4684 (97.8) | 26.6 (7.1) |
| Non-smoker | 2128 (39.0) | 1912 (39.9) | 216 (32.5) |
| Active-smoker | 486 (8.9) | 446 (9.3) | 40 (6.0) |
| Ex-smoker | 2714 (49.8) | 2326 (48.6) | 388 (58.4) |
| Co-morbidity * | 3001 (55.1) | 2454 (51.3) | 547 (82.4) |
| All medications ** | 2452 (45.0) | 1980 (41.4) | 472 (71.1) |

\* Includes hypertension, coronary heart disease, diabetes (type 1 or type 2), chronic kidney disease stage 3-5, asthma, COPD, immunocompromised
\*\* Includes antihypertensive medication, lipid lowering medication, hypoglycaemic medication, inhalers, immunosuppressants

### HRT use and all-cause mortality in COVID-19

HRT use was associated with a lower likelihood of all-cause mortality in COVID-19 within unadjusted models (OR 0.15, 95%□CI 0.06 to 0.37) and adjusted models (OR 0.22, 95%□CI 0.05 to 0.94). We also observed that all-cause mortality risk was higher in COVID-19 amongst women who were older, underweight, from larger households, with hypertension, or on immunosuppressants. For those with asthma, this association seemed to be protective as women on HRT had a significantly lower odds of all-cause mortality (OR 0.58, 95%□CI 0.42 to 0.81). These results are shown in table 3 below.

**Table 3:**
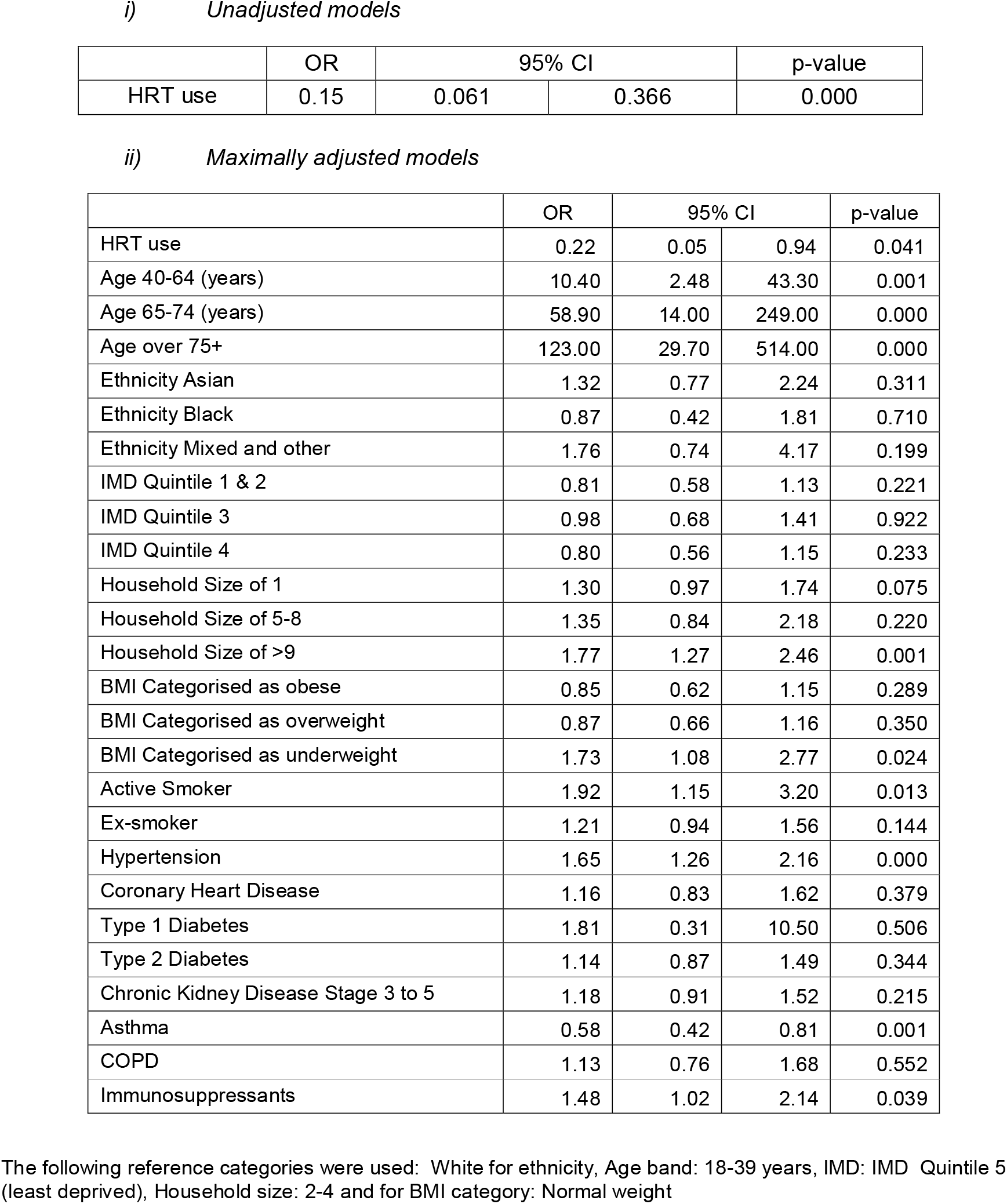
Association between HRT use and the likelihood of death in women with COVID-19 (n= 5451)

### COCP use and all-cause mortality in COVID-19

We had intended to examine COCP as an exposure but as there were no reported events for the outcome of interest (all-cause mortality) in women prescribed COCPs. Accordingly, we were unable to examine COCP use.

## DISCUSSION

### Main findings

In this cohort of 5451 women with COVID-19 who were followed up in the first six months of the pandemic, HRT use was associated with a lower likelihood of all-cause mortality.

### Strengths and limitation

A major strength of this study is the use of a population-based cohort from 465 practices across England representing wide coverage with a denominator population of 3.6 million people. This included heterogeneity in sociodemographic and clinical variables. The data used is of high quality and completeness with twice-weekly updates that are also used by Public Health England to monitor the current and previous pandemics.[9] The availability of wide-ranging and precise data means that we were able to adjust for several confounders, although residual confounding is still possible. We considered both laboratory-confirmed and clinically probable cases as a single cohort due to the national inconsistency in testing availability. It is plausible that not all those with clinically probable cases had SARS-CoV-2. Recent work in Oxford RCGP database suggests that outcomes are similar in those with clinically probable and laboratory-confirmed cases.[17] Serology testing, if available nationally in the future could also be helpful. Our cohort is likely to reflect women with more severe COVID-19 symptoms who went for testing or made contact with a general practice for review. If asymptomatic or with milder symptoms, they may not have sought health advice and will not be captured in this cohort. In terms of the exposure, we examined medications based on prescriptions within the last 6 months rather than dispensed medications so there could be some over-ascertainment of exposure to oestrogens. Further, as oestrogen was highlighted as having a role in COVID-19 reasonably early in the pandemic, it is possible that some women may have stopped taking their medications before contracting the infection. Our study did not examine the type of preparation or dose of HRT. Nor did we investigate the duration of medication use and our follow-up period was short at less than six months. This might be important in oestrogen related immune responses where longer exposures to hormones could be significant.[2] We used all-cause mortality as our outcome, and some deaths may therefore be unrelated to COVID-19. There was substantial confusion about the classification of COVID-19 mortality in the early part of the pandemic including changes in Government guidance as the pandemic progressed. COVID-19 specific mortality was variable and as a new code, it may not have been widely used in primary care records. All-cause mortality is likely to be a more reliable measure especially in the early part of the pandemic in which our study is set.

### Interpretation

To our knowledge, this is the first study to examine the use of exogenous oestrogens through HRT concerning all-cause mortality amongst women with COVID-19. Previous studies report lower rates of severe COVID-19 complications amongst women compared to men, and our findings give weight to current hypotheses suggesting that oestrogen may confer a protective effect against COVID-19. [7,8] [4,5] This is consistent with the findings of the COVID Symptom Study which is the largest observational study to date including 152,637 women for menopause status. [7] Their findings across the cohort suggests that higher oestrogen levels may protect against COVID-19. The mechanism to explain this may be through increased cellular and humoral immune responses in females with higher oestrogen levels. Recent evidence suggests that females have a higher level and faster generation of serum SARS-CoV-2 IgG antibody compared to males.[18] Higher oestrogen levels may also be able to better promote direct anti-viral activity of T-cells and modulate the uncontrolled immune response (cytokine storm) that has been observed in those with respiratory failure due to COVID-19. [4,5] Immune responses and oestrogen level decrease with age which might explain why previous studies and our results show a greater likelihood of worse outcomes in females with increasing age. [1,7] However, amongst women on HRT with exogenous oestrogen the risk of all-cause mortality are reduced but still do not reach that of younger females presumably related to the direct effect of ageing on the immune system, and the increased number of morbidities acquired with age.[19] In the COVID Symptom Study described earlier, the association between HRT and COVID-19 hospitalisation in 17 798 women were variable and the authors do not show that HRT lowers the risk of hospitalisation but they do not report on mortality. [7] These differences might be explained by variations in HRT preparations, doses and duration which were not examined and as described above might be important in oestrogen led immune responses.[2] Other explanations may relate to differences in adjusted covariates which were limited to age, smoking and BMI in their study. As the pandemic progresses and a greater understanding of the virus emerges, it is necessary to consider additional covariates such as household size and co-morbidities which we included.[1] Our results show that increased age, co-morbidities, extreme BMI and immunosuppressants were all significantly associated with an increased likelihood of death amongst women with COVID-19; this is consistent with several recent reports.[20,21] There is some uncertainty in the literature about the role of asthma in the severity of COVID-19 outcome but we observed that being on HRT was associated with a significantly lower risk of mortality (OR 0.58, 95%□CI 0.42 to 0.81), suggesting that perhaps oestrogen is protective. However, these women with asthma are likely to also have been on asthma medication such as steroids which could contribute to some of the observed associations. [22]

## Conclusions

We found that HRT use in COVID-19 infection was associated with a lower likelihood of death. Women on these medications should be encouraged to continue to take HRT during the pandemic. Further work is needed to explore the effect of variations in HRT doses, preparations and duration on COVID-19 complications. Additional research is also required in larger cohorts to examine the association been COCP and mortality in COVID-19.

## Data Availability

Data are available by direct request to the database https://www.rcgp.org.uk/clinical-and-research/our-programmes/research-and-surveillance-centre.aspx

## AUTHOR CONTRIBUTION

HDM designed the study, wrote the first draft and edited subsequent versions of the manuscript. WH contributed to study design, led the data analysis and revised the manuscript. MJ provided advice on statistical methods. MF and Sdel contributed to the study design and revised the paper. Sdel also provided expertise on the RCGP RSC database.

## ETHICAL APPROVAL

This study received approval from the Oxford-RCGP RSC study approval committee (RSC_0920) and the University of Southampton Research Ethics committee (56309) on the 6th May 2020.

